# First month of the epidemic caused by COVID-19 in Italy: current status and real-time outbreak development forecast

**DOI:** 10.1101/2020.03.26.20044628

**Authors:** Rosario Megna

## Abstract

**Background:** The first outbreaks of COVID-19 in Italy occurred during the second half of February 2020 in some areas in the North of the country. Due to the high contagiousness of the infection, further spread by asymptomatic people, Italy has become in a few weeks the country with the greatest number of infected people after China. The large number of severe cases among infected people in Italy led to the hospitalization of thousands of patients, with a heavy burden on the National Health Service.

**Methods:** We analyzed data provided daily by Italian Authorities for the period from 24 February 2020 to 26 March 2020. Considering such information, we developed a forecast model in real-time, based on the cumulative logistic distribution. We then produced an estimate of the overall number of potentially infected individuals and epidemic duration at a national and Regional level, for the most affected Regions.

**Results:** We reported the daily distribution of performed swabs and confirmed cases, and the cumulative distribution of confirmed cases, of patients quarantined at home (42%), hospitalized in non-intensive care (31%), recovered or discharged (13%), deceased (10%), and hospitalized in intensive care (4%). The forecast model estimated a number of infected persons for Italy of 115,000 about, and a duration of the epidemic not less than 2 months.

**Conclusions:** Once month after the first outbreaks there seems to be the first signs of a decrease in the number of infections, showing that we could be now facing the descending phase of the epidemic. The forecast obtained thanks to our model could be used by decision-makers to implement coordinative and collaborative efforts in order to control the epidemic.

## Introduction

The outbreak of the novel Coronavirus called COVID-19 (or SARS-CoV-2), which originated at the end of December 2019 in the city of Wuhan, in the Hubei Province, China [1], is having a dramatic global evolution, and was recently classified as a pandemic by the World Health Organization (WHO) on 11 March 2020 [2]. The disease, which can be diagnosed through the use of a nasopharyngeal swab, under the most severe forms can lead to bilateral pneumonia [3] which can be lethal especially in elderly patients with comorbidities [4]. The first outbreaks of COVID-19 in Italy occurred during the second half of February 2020 in some areas in the North of the country. Despite the drastic restrictions imposed by the Italian Government in those areas (DL n. 6, 23 February 2020) [5], several other outbreaks began in other areas of northern Italy, forcing the Authorities to extend the previously adopted restrictions to the entire national territory (DPCM of 3 March 2020 et seq.) [5]. Due to the high contagiousness of the infection, further spread by asymptomatic people [4], in a few weeks Italy has become the country with the greatest number of infected people after China [6]. The large number of severe cases among infected people in Italy led to the hospitalization of thousands of patients [7,8], with a heavy burden on the National Health Service. In particular, the most affected Regions are Lombardy and Emilia Romagna, with more than half of the total cases. One month after the beginning of the epidemic in Italy we report the situation and propose a forecast model in real-time to estimate its evolution.

## Materials and Methods

Data on COVID-19 used in our analysis are daily updates from the Italian Ministry of Health managed by the Civil Protection Department [7,8]. A report is released at 5:00 or 6:00 pm (CET), on the basis of information provided by national and Regional Local Authorities. The most relevant variable is the number of confirmed cases. The other derivate variables to be considered are the number of hospitalized patients (in intensive or non-intensive care), individuals quarantined at home, patients who recovered or were discharged, and the number total deaths.

We analyzed data used in this study using the R software, version 3.6.3 (R Foundation for Statistical Computing, Vienna, Austria). Continuous variables were expressed as mean ± standard deviation (SD) and categorical variables as percentages, while differences between groups were evaluated by χ^2^.

We developed a forecast model in real-time, based on the cumulative logistic distribution [9]. The equation used is the following:

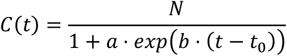

where *C*(*t*) is the cumulative incidence on day *t, N* is the cumulative incidence at the end of the epidemic, *a* is the parameter that governs the flexibility of the curve, *b* is the logistic growth rate, and *t*_0_ is the initial day of the epidemic analysis (24 Feb 2020). In order to determine the three parameters of the curve, we developed an algorithm based on the maximum statistical significance of *a* and *b*, according to their p-values (smaller values of p-value indicate greater significance), and varying *N*. Nonlinear least squares (nls) function of R was used, normalizing *C*(*t*)→*C*(*t,N*=1)≡*C*′.The steps of the algorithm are the following:

~~~
***for*** *n (from min_n to max_n, step delta_n)*
       *nls (C*′ *(a(n), b(n)))*
       ***if*** *(p-value(a (n)) < p-value(a(n-1))* ***and*** *(p-value(a (n)) < p-value(a(n-1))*
       ***then***
            ***continue***
       ***else***
       *a = a(n-1); b = b(n-1); N = n-1*
~~~

with min_n = 10000, max_n = 150000, and delta_n = 1000 for the National evaluation and min_n = 5000, max_n = 70000, and delta_n = 500 for the Regional evaluations. At the end of the last cycle, the algorithm provides parameters for best-fit. In order assess the 95% confidence interval (CI) of the fit values, 1000 bootstrap resampling were computed, through the IPEC package of R. The graphics were obtained using the ggplot2 package of R.

## Results

Figure 1 shows the daily distribution of performed swabs and confirmed cases. The percentages on the bars are related to the ratio between the two variables (8.6% ± 8.1%). Until 26 March 2020, the total number of performed swabs is 361,060. In Figure 2 we report the cumulative distribution of confirmed cases, of patients quarantined at home, hospitalized in non-intensive care, recovered or discharged, deceased, and hospitalized in intensive care. In Table 1 is reported the number of confirmed cases and the patient categories (expressed in percentage) for at a National and Regional (Lombardy and Emilia Romagna) level. The confirmed cases are greater than 80,000, while the number of deceased patients is significantly greater than the Chinese [8] (P < 0.001). In table 2 we summarized the forecasted method results for at a National and Regional (Lombardy and Emilia Romagna) level. In figure 3 we depict the cumulative logistic curve obtained by the forecast model in real-time for the national overview.

**Table 1.** Confirmed cases and patients categories (expressed in percentage).

|  |  | **Patients categories** |  |  |  |  |
| --- | --- | --- | --- | --- | --- | --- |
|  | **Confirmed cases**<br>**(26 March 2020)** | **Quarantined at**<br>**home** | **Hospitalized in**<br>**non-intensive care** | **Recovered or**<br>**discharged** | **Deceased** | **Hospitalized in**<br>**intensive care** |
| **Italy** | 80,539 | 42% | 31% | 13% | 10% | 4% |
| **Lombardy** | 34,889 | 29% | 31% | 22% | 14% | 4% |
| **Emilia**<br>**Romagna** | 10,816 | 48% | 31% | 7% | 11% | 3% |

**Table 2.** Best-fit parameters and relevant dates obtained by the forecasted model for the epidemic caused by COVID-19 in Italy. Time such that C(t)=0.5 is related to the curve inflection point, i.e. the descending phase of the epidemic has started.

| Forecast | Best-fit parameters |  |  | Relevant dates<br>(number of days since the outbreak began) |  |
| --- | --- | --- | --- | --- | --- |
| | a (p-value) | b (p-value) | N (CI) | $C(t)=0.5$ | $C(t)=0.99$ |
| **Italy** | 229.0 ($<10^{-5}$) | -0.1939 ($<10^{-5}$) | 117,000 (112,000-122,000) | 22 March (28) | 27 April (64) |
| **Lombardy** | 133.4 ($<10^{-5}$) | -0.1870 ($<10^{-5}$) | 46,000 (44,000-47,500) | 20 March (26) | 14 April (51) |
| **Emilia**<br>**Romagna** | 198.2 ($<10^{-5}$) | -0.1679 ($<10^{-5}$) | 21,000 (20,000-22,000) | 26 March (32) | 22 April (59) |

**Figure 1.**
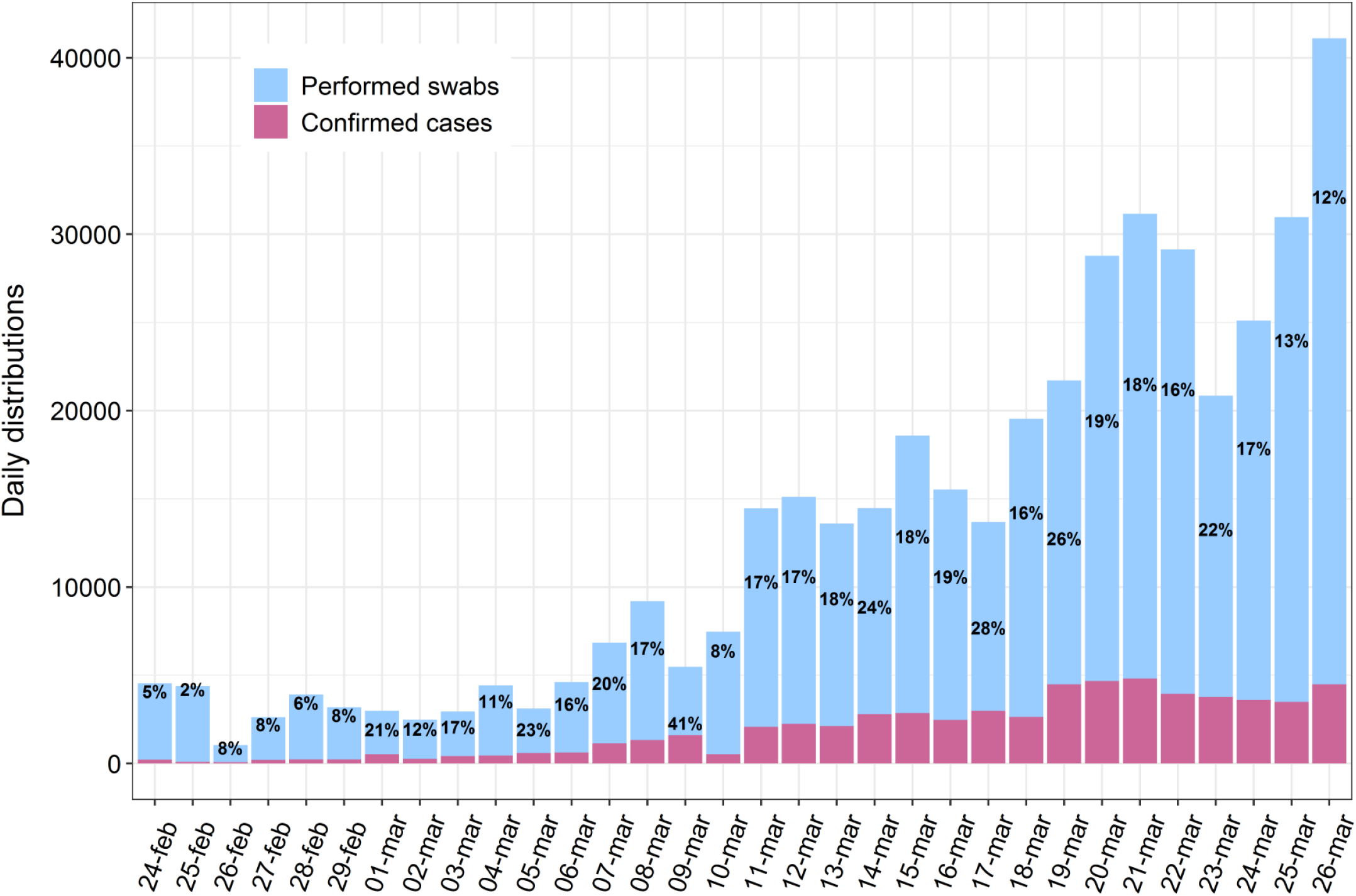
Daily distribution of the performed swabs and confirmed cases. The percentages are related to the ratio between confirmed cases and performed swabs.

**Figure 2.**
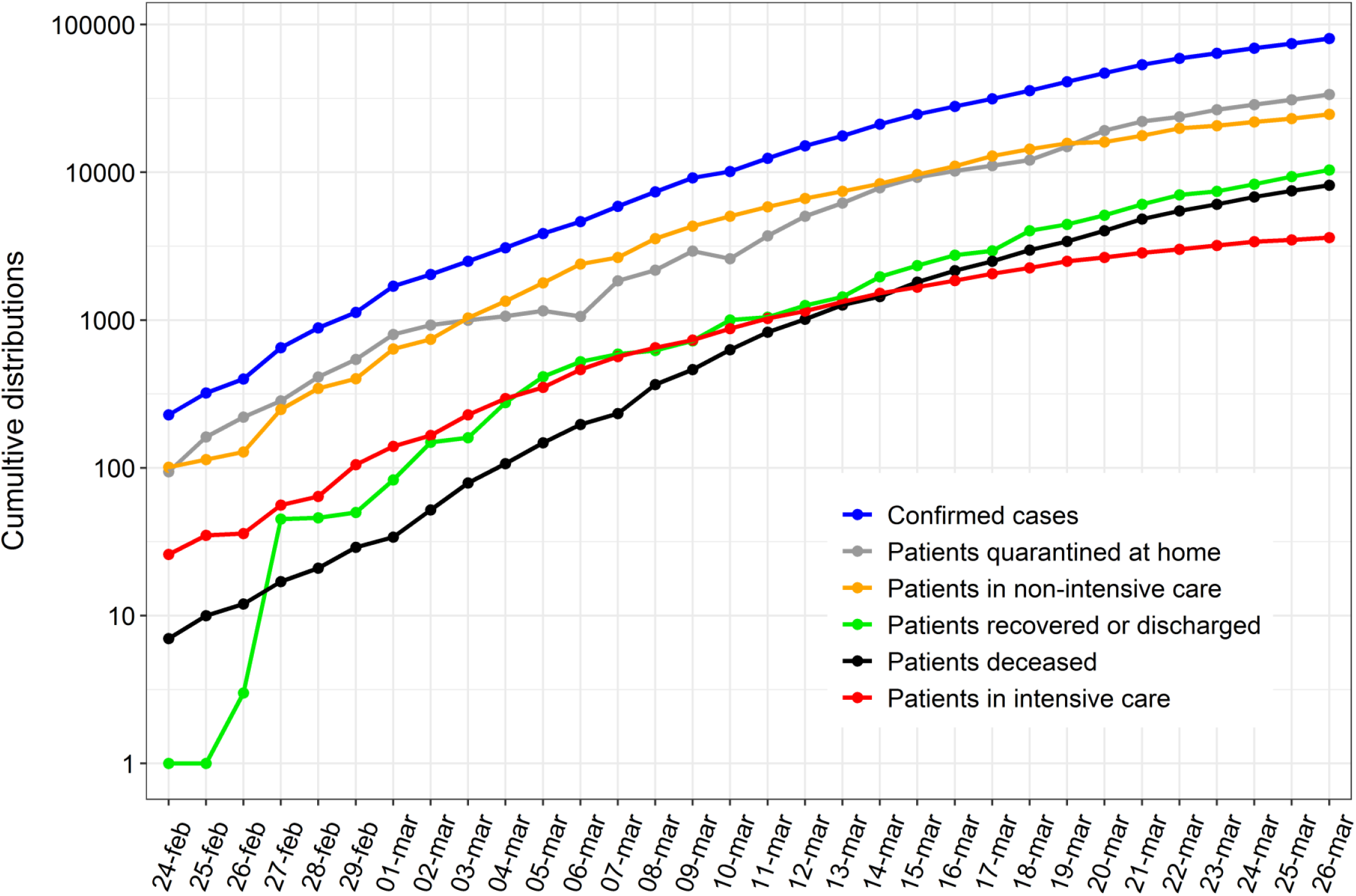
Cumulative distribution of the confirmed cases and patients categories.

**Figure 3.**
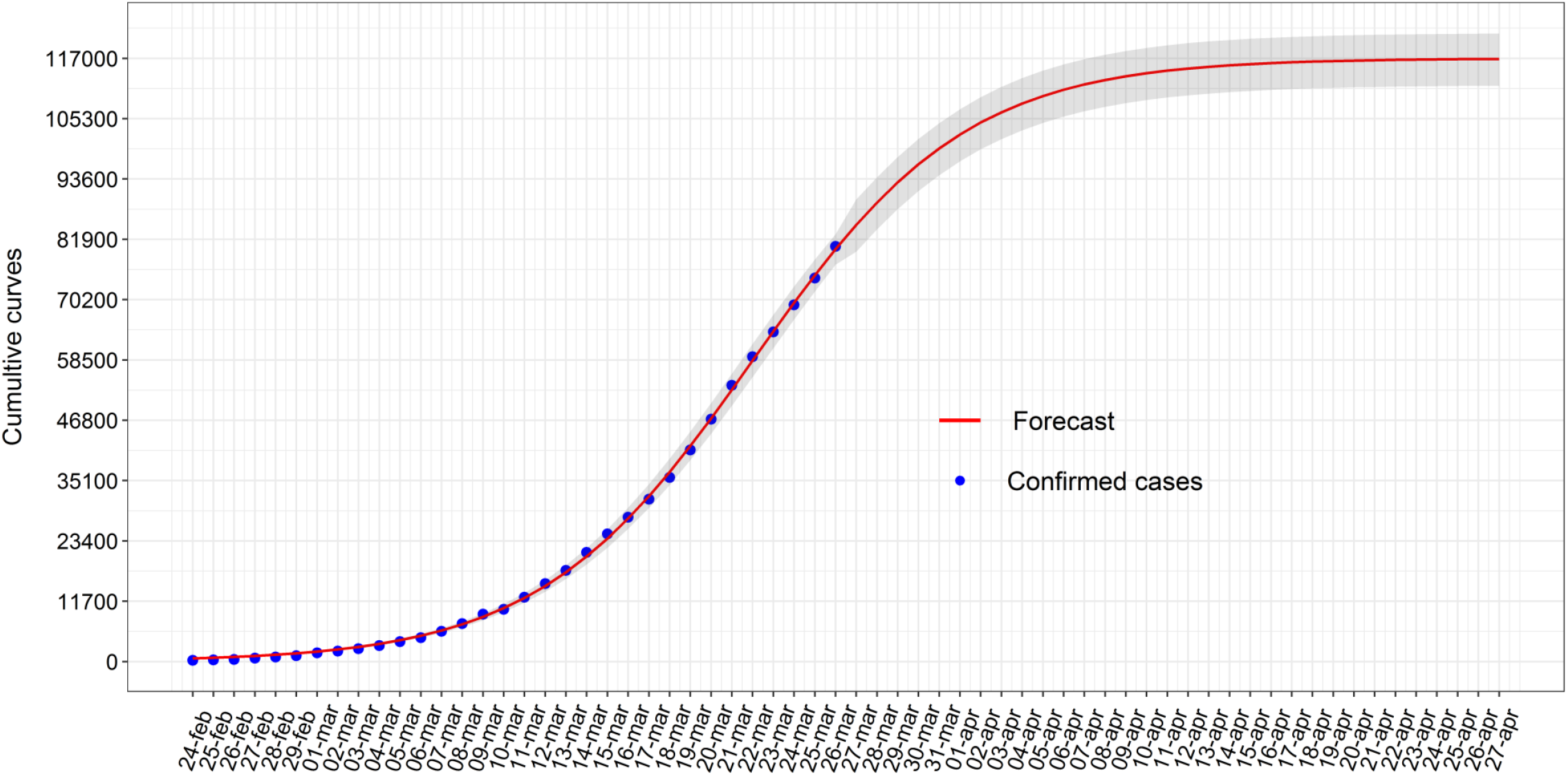
Cumulative curve obtained by the forecasted model of the epidemic in Italy in real-time. Predictions are represented by the red line, with the gray area to indicate 95% CI. The blue points represent the confirmed cases.

## Discussion

One month after the outbreak in Italy the situation remains complicated. Despite the high number of performed swabs as compared to the confirmed cases, the epidemic has been growing with a very high rate. COVID-19 is proving to have a high capacity for infection, probably reinforced by asymptomatic people, who represent a real danger for elderly and fragile individuals. In particular, the disease is showing to be lethal for the elderly (96% in patients aged ≥ 60) and men (71%) [10]. On the date we finalized this article (26 March 2020), the trend of daily distribution of confirmed cases seems to show an initial decline of the growth of the epidemic. The total number of confirmed cases will eventually exceed those that occurred in China. The forecast model in real-time indicates a total number of national cases greater than 110,000 patients, with a figure of approximately 46,000 in Lombardy only. In addition, duration of the epidemic was estimated of 2 months about. Since the theoretical cumulative curve has an asymptotic pattern (i.e. the maximum value is achieved for the t time towards infinite), considering 99% of the time from the beginning of the outbreak is a convention. Therefore, if instead of 99% of time we considered 99,9%, the overall number of days estimated for the epidemic to come to an end increases by 20% (i.e. ten more days need to be added to the calculation of time). Moreover, several factors could affect the total number of cases and the duration of the epidemic. For example, a contribution to the spread of outbreaks in southern Italy was caused by the movement of students and workers from Northern to Southern Italy following the first governmental restrictions. On the other hand, more stringent restrictions imposed later on by the Government could lower the expected number of total cases and reduce the number of days towards the end of the epidemic. Another important factor is related to possible mutations of the novel Coronavirus, which could have a positive or negative outcome on the trend of the epidemic.

It is also necessary to consider the intrinsic limitations of this study. First of all, data was not always updated on a daily basis by each Regional Authority. Moreover, we have to consider that the number of infected people is underestimated, since there are many undetected asymptomatic individuals. In addition to this, many individuals died without the possibility of checking if they were actually infected and therefore not recorded as such. Finally, the factors that determine the trend of the epidemic could change without respecting the symmetry of the forecasted model.

## Conclusions

The epidemic caused by COVID-19 in Italy is having a dramatic evolution in terms of confirmed cases, hospitalized and deceased patients. Once month after the first outbreaks there seems to be the first signs of a decrease in the number of infections, showing that we could be now facing the descending phase of the epidemic. The model presented in this article fits well with the data, therefore it is expected to be reliable in predicting the evolution of the epidemic. The forecast could be applied by decision-makers to take coordinative and collaborative efforts to control the epidemic.

## Data Availability

Data used are available online

## Ethics approval and consent to participate

Not applicable.

